# Public perception of organ donation and transplantation policies in Southern Spain

**DOI:** 10.1101/2021.09.17.21263724

**Authors:** Gonzalo Díaz-Cobacho, Maite Cruz-Piqueras, Janet Delgado, Joaquín Hortal-Carmona, M. Victoria Martínez-López, Alberto Molina-Pérez, Álvaro Padilla-Pozo, Julia Ranchal-Romero, David Rodríguez-Arias

## Abstract

**Background:** This research explores how public awareness and attitudes towards donation and transplantation policies may contribute to Spain’s success in cadaveric organ donation.

**Materials and Methods:** A representative sample of 813 people residing in Andalusia (Southern Spain) were surveyed by telephone or via Internet between October and December 2018.

**Results:** Most participants trust Spain’s donation and transplantation system (93%) and wish to donate their organs after death (76%). Among donors, a majority have expressed their consent (59%), while few non-donors have expressed their refusal (14%). Only a minority is aware of the presumed consent system in force (28%) and feel sufficiently informed regarding the requirements needed to be an organ donor (16%). Participants mainly consider that relatives should represent the deceased’s preferences and be consulted when the deceased’s wishes are unknown, as is the case in Spain.

**Conclusions:** Public trust in the transplant system may contribute to Spain’s high performance in organ donation. High levels of societal support towards organ donation and transplantation do not correspond in Spain with similar levels of public awareness of donation and transplantation policies.

## Introduction

The scarcity of organs for transplantation is a global problem that is internationally addressed by using different strategies, such as increased investment in healthcare infrastructure (e.g. staffing transplantation coordinators at hospital facilities, and different systems of economic remuneration for healthcare professionals involved in the donation and transplantation process), switching to an opt-out consent model for organ recovery[1,2] boosting public information through donation campaigns,[3] and different schemes for organ allocation.[4,5]

Spain has the highest rate of organ donors worldwide with 48.9 donors per million people (henceforth pmp) in 2019, well above international averages.[6] The country has a presumed consent model in which the family is systematically asked about the deceased’s preferences, and where family opposition is always respected.[7] Organ recovery relies on brain death donation (70%) and two types of donation after circulatory death (DCD): controlled DCD and uncontrolled DCD (25%).[7] The organ donation system is structured in a multi-level transplant coordination system with decades-long investment in specific infrastructures designed for removing obstacles to donation. This includes staffing hospitals with transplant coordinators –mainly intensive care doctors– endowed with specific responsibilities to enable organ recovery through identification and inclusion of the donation option in end-of-life procedure.[8]

Health professionals involved in organ donation may receive, on top of their regular salary, retribution for their availability during organ donation on-call shifts. In some regions, including Andalusia, their activity and performance may also be remunerated.[8] A centralized organ distribution system guarantees countrywide access to urgently needed organs while simultaneously prioritizing local allocation.[9] Finally, emphasis is made on the media as a form of swaying public opinion in favor of organ donation through proactive messaging, an ongoing effort to provide information, and case-by-case management of information crises.[9]

The implementation of similar policies has contributed to raise organ donation rates in other regions, but remaining differences in organ availability between Spain and other countries suggest that cultural factors –rather than only structural ones– might be playing a role too.

The success of any transplantation system is based on people’s willingness to participate and public attitudes favorable to donation. Lower levels of public trust in health institutions can decrease organ supply and harm overall transplant performance. Despite some studies on the factors that influence people’s willingness to donate in Spain, including their views on the consent model, we still don’t know the extent to which Spaniards’ attitudes toward donation may underlie the country’s success. [10–13]

In this paper, we surveyed a representative sample of the population in Andalusia, Spain’s southernmost region, to assess their awareness and general attitudes regarding organ donation and transplantation –i.e. trust and willingness to donate–, and their views on customary donation and transplantation policies: a) presumed consent for organ retrieval; b) allowing the families of the deceased to make decisions regarding organ removal; c) allowing donation after brain death as well as donation after circulatory death; and e) health professionals’ retribution based on donation activity. Finally, we explored their preferred criteria for organ allocation and their view on the notion that organ donation can facilitate family grieving.

## Materials and Methods

Between October and December 2018, we carried out a survey coinciding with the 7th Edition of the Citizens’ Panel for Social Research in Andalusia (PACIS/EP-1807). The region of Andalusia is especially interesting to study because its organ donation rate is higher than the national average (51.5 donors ppm) and it is both the most populated region in the country (8.5□M inhabitants) and one of the most diverse with 45% of its population living in urban centers (such as Seville, Malaga, Cordoba) and 17% in rural areas.

The sample, stratified by gender and age, was drawn from 1,929 out of the 3,700 people belonging to the PACIS Panel (its methodology is available at<u>(3)</u>). As a result, our final sample was formed by 813 people aged 18 and older. We calibrated the sample using the raking method to the gender, age, educational level and municipal population size variables with figures from the total Andalusian population as a reference. The maximum sample error of the study is +/–3.5%. The questionnaire (see Supplementary File), containing a total of 22 questions related to organ donation, was implemented through telephone surveys (46%) and online (54%), and lasted an average of 19 minutes. Participants were compensated with 5 euros, which they could either keep or donate to a non-governmental organization.

All procedures performed in this study were carried out in accordance with the European Charter of Fundamental Rights and with the Declaration of Helsinki and its later amendments. The study protocol was reviewed by the Coordinating Committee on Biomedical Research Ethics of Andalusia (PEIBA 2521-N-20), which waived full review for this type of study. Written informed consent was obtained from all individual participants prior to participation.

The average age of the sample was 48 years; 52% of participants were women; 75% had high school or further education; 68% considered themselves religious and 26% stated they were quite or very observant; 33% stated they had had some sort of close experience with organ donation or transplantation concerning a family member or close friend. Half of those surveyed chose to donate their reward for answering the questionnaire to an NGO (Table 1).

**Table 1:**
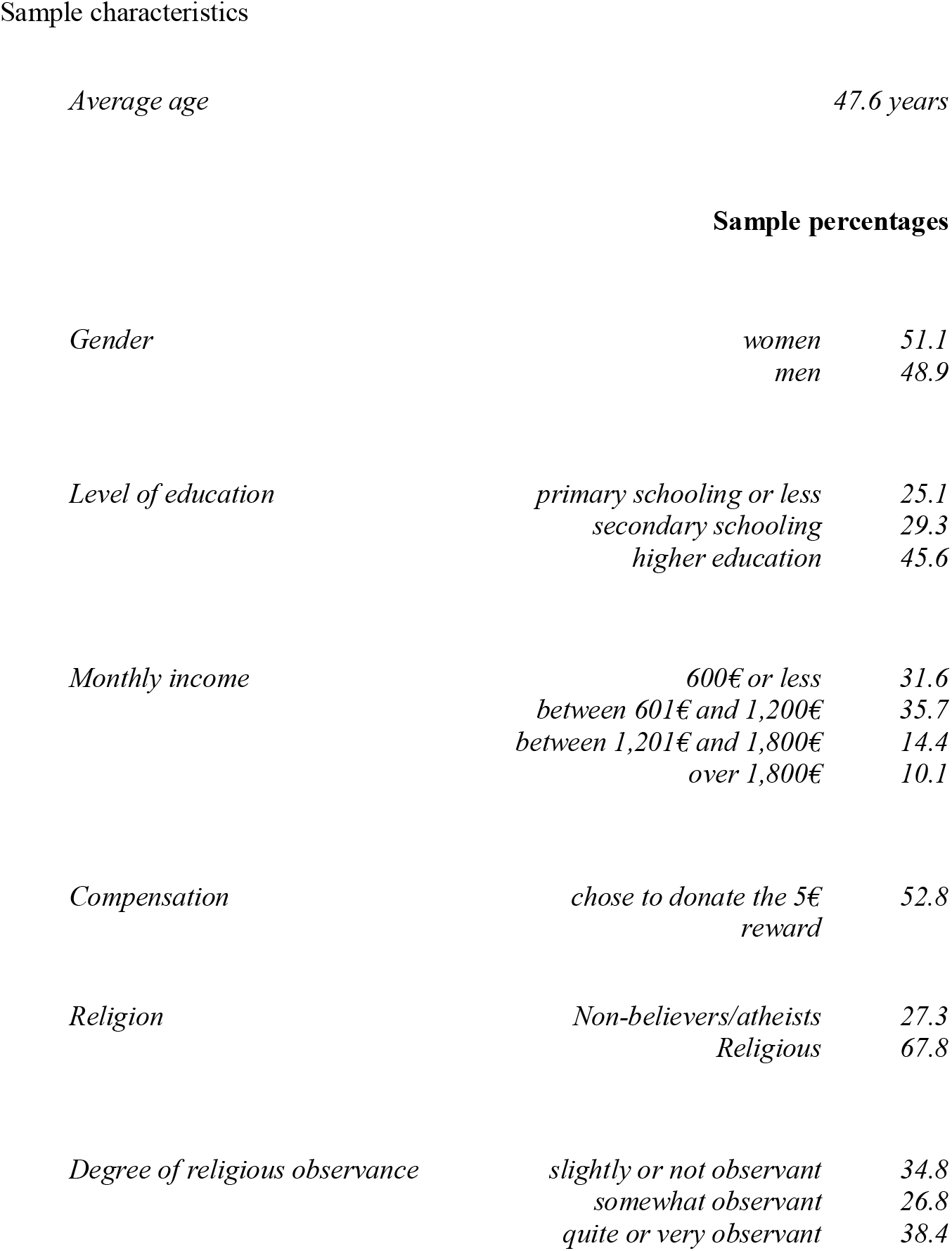
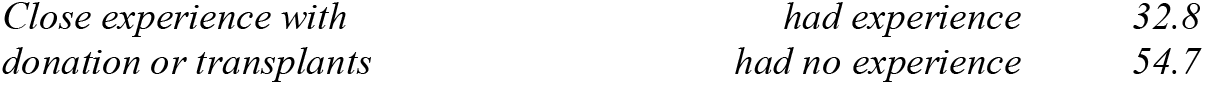
Sociodemographic data.

|  |  |
| --- | --- |
| *Average age* | *47.6 years* |

Sample percentages
|  |  |  |
| --- | --- | --- |
| *Gender* | *women* | *51.1* |
|  | *men* | *48.9* |
| *Level of education* | *primary schooling or less* | *25.1* |
|  | *secondary schooling* | *29.3* |
|  | *higher education* | *45.6* |
| *Monthly income* | *600€ or less* | *31.6* |
|  | *between 601€ and 1,200€* | *35.7* |
|  | *between 1,201€ and 1,800€* | *14.4* |
|  | *over 1,800€* | *10.1* |
| *Compensation* | *chose to donate the 5€ reward* | *52.8* |
| *Religion* | *Non-believers/atheists* | *27.3* |
|  | *Religious* | *67.8* |
| *Degree of religious observance* | *slightly or not observant* | *34.8* |
|  | *somewhat observant* | *26.8* |
|  | *quite or very observant* | *38.4* |
| *Close experience with*<br>*donation or transplants* | *had experience* | 32.8 |
|  | *had no experience* | 54.7 |

The survey addressed questions on the following topics: 1. general attitudes toward donation and transplantation (i.e. trust in the healthcare and donation/transplantation systems, and willingness to donate); 2. awareness of the criteria for determining death and willingness to donate in three types of cadaveric donation circumstances; 3. awareness and attitudes regarding the consent model for donation and the role of the family, 4. attitudes on allocation criteria, 5. opinions on health professionals’ retribution based on donation activity, and 6. views on family grieving. Topics 2 and 4 were preceded by a briefing on cases regarding cadaveric donation and organ allocation options.

The statistical analysis of the data was descriptive and exploratory. First, we analyzed the relative frequencies of the most relevant questions for this study. Second, we used contingency tables to explore the role of gender, age, level of education, religion and political affiliation on those questions using Pearons’s Chi-squared tests. Only significant correlations with a p-value ⍰ .1 were considered.

## Results

A majority of the population surveyed displays trust in the public healthcare system (79%), and especially in the donation and transplantation system (93%), which most (59%) consider to be transparent. Elderly people and those who classify themselves as politically left-wing tend to exhibit greater trust in the transplantation system. Three out of four people surveyed state that they would donate their organs after death and 62% would authorize organ retrieval for a family member whose wish to be or not to be a donor is unknown. A majority (59%) of those who wish to be donors but a minority (14%) of those who refuse donation have made a verbal or written record of their preference. Self-defined politically right-wing individuals and self-defined Catholics do significantly (p.<0.05) oppose donation more than other groups (see Supplementary Table 1-4).

When presented with prototypical scenarios for the three situations in which cadaveric donation occurs, a minority of those surveyed recognize organ recovery as legal in cases of brain death (19%), controlled DCD (22%) and uncontrolled DCD (21%) (Table 2). A similar proportion assert that they would agree to have their organs removed for transplantation purposes in each of the three aforementioned situations.

**Table 2:**
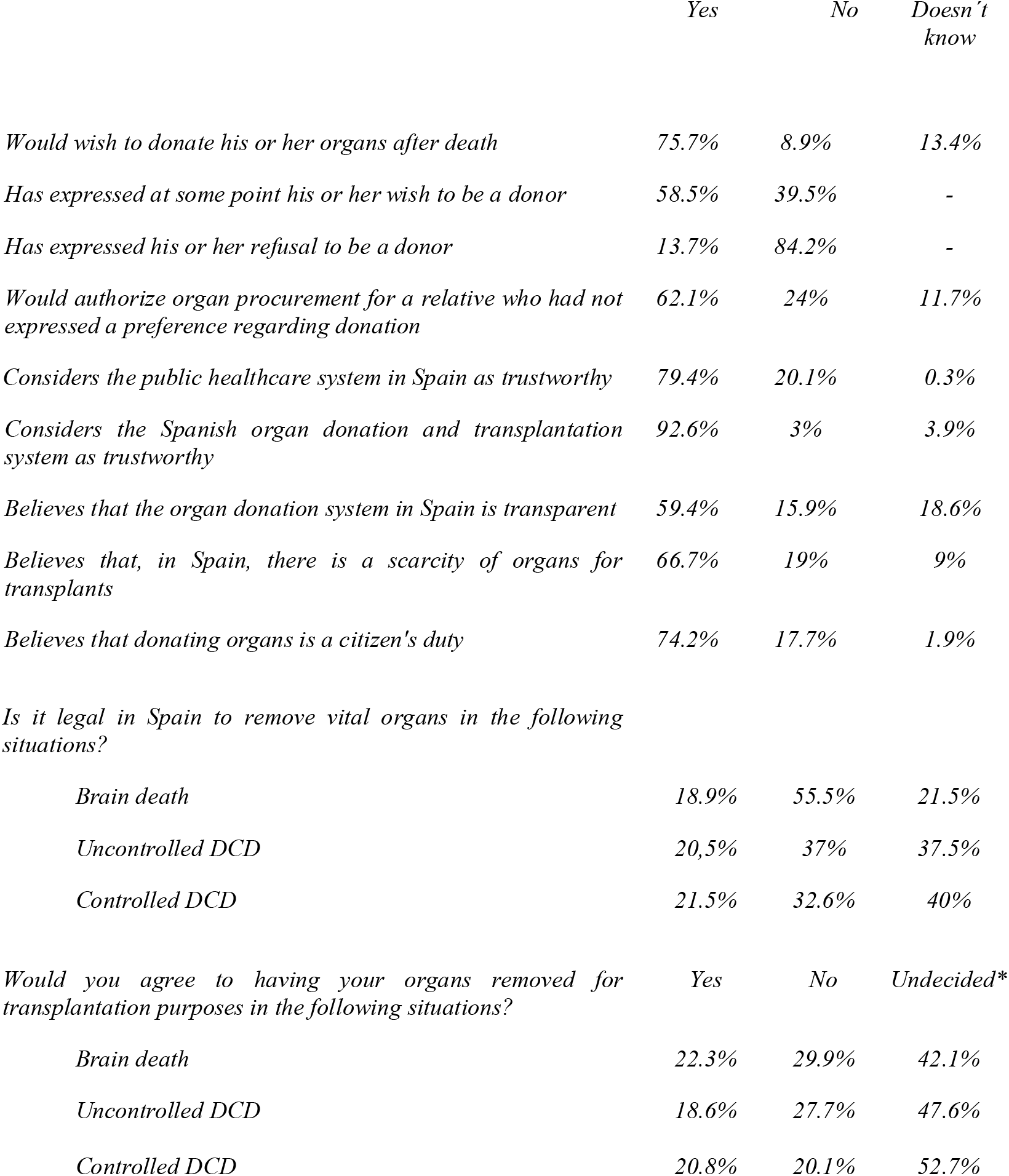
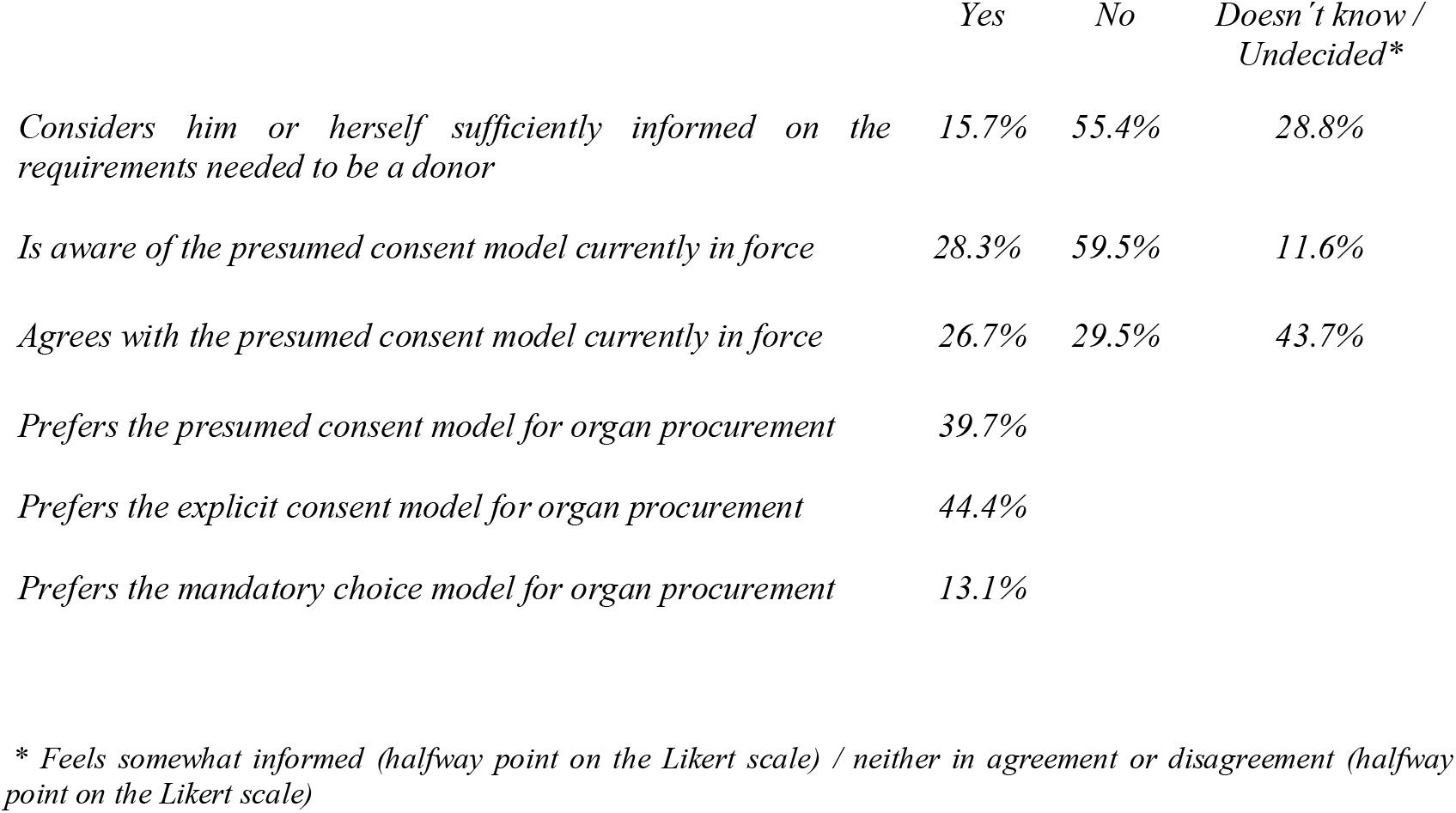
Attitudes and awareness regarding donation and transplantation, criteria for determining death, and consent models for donation.

Over half of respondents feel somewhat or completely uninformed on the requirements needed to be an organ donor (Table 2). A majority erroneously believe that Spain is governed by an opt-in system or recognize to be unaware of the consent model currently in force for organ recovery in Spain. Participants’ opinions on the presumed consent model are divided: 30% oppose it, 27% support it and the remaining 44% are undecided on the subject. When asked to choose from among three different models, those surveyed are divided between 44% who side with opt-in, 40% who prefer opt-out and 13% who choose the mandatory choice model. Non-believers or atheists are more inclined to the opt-out model than are Catholics. They also are more familiar with the current model in force in Spain than Catholics and those who profess other beliefs. People who identify as ideologically more left-wing mostly prefer an opt-out consent model, as opposed to those in the center or on the right-wing who mostly support the opt-in model (see Supplementary Table 1-4).

While making decisions on organ donation, three distinct situations may arise: the deceased expressly consented to donate, expressly refused to donate, or failed to express any preference. During the donation interview, the preferences of the deceased are systematically explored through the family, which is nevertheless allowed to make the ultimate decision to authorize or oppose organ recovery. Most respondents consider that, whenever the deceased had expressed a preference, either in favor or against donation, “the medical team should ask the family whether or not the person had changed their opinion, because one should always respect the deceased’s most recent wishes” (Table 3a). When the deceased had not expressed any preference, most respondents consider that “the family should convey what they believe to have been the deceased’s wishes”.

**Table 3:**
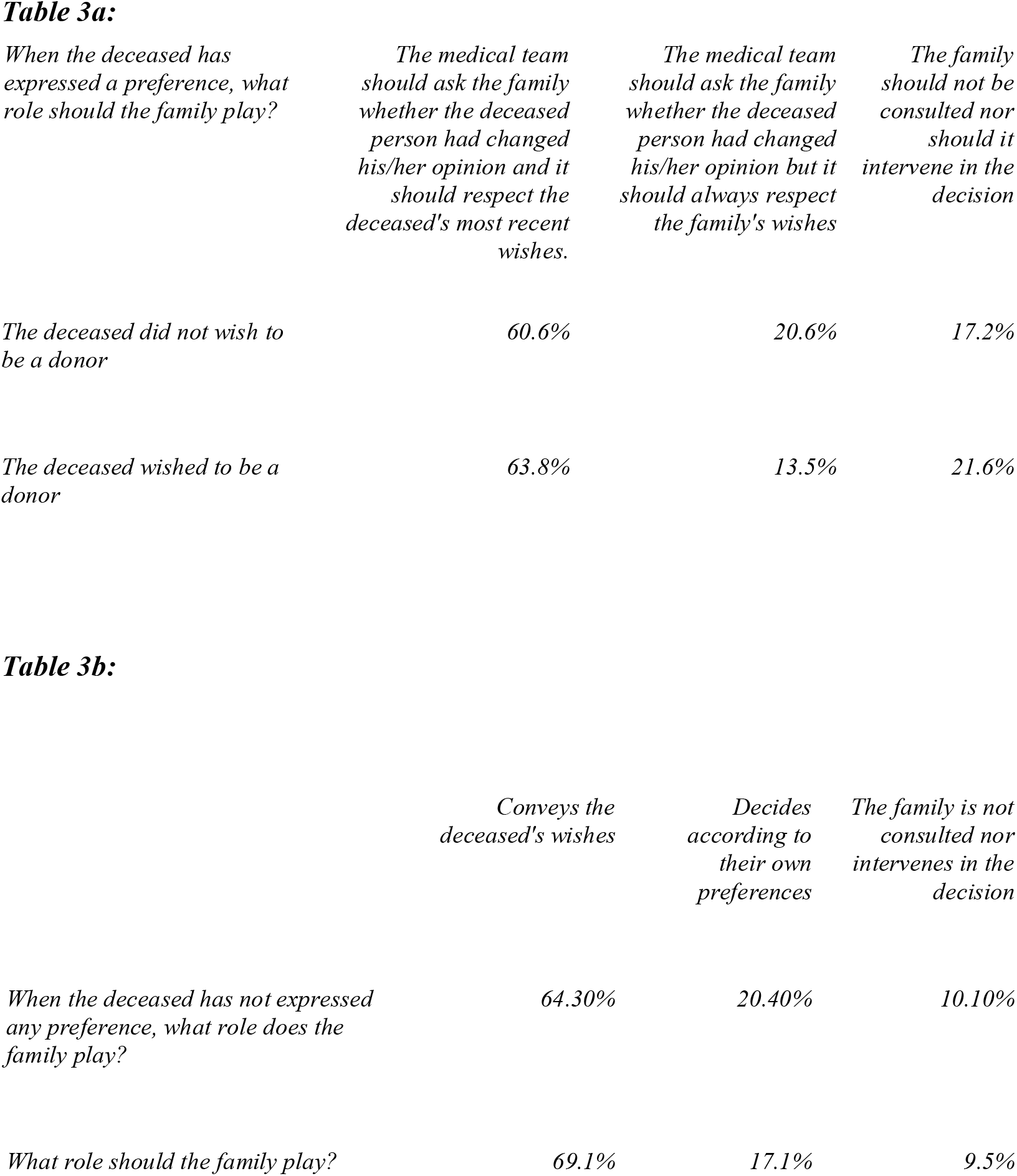
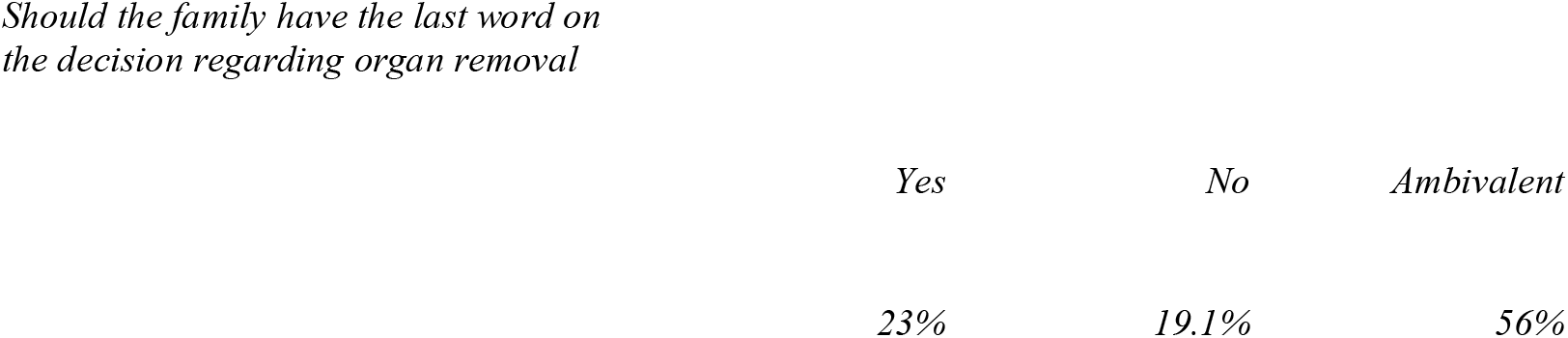
Preferred role of the family and deceased expressed vs non-expressed preferences.

In clinical practice, families are systematically asked to make a decision when the deceased has not. We wished to further explore the population’s awareness of this policy and learn its opinion on the subject. One out of ten surveyed erroneously think that, in Spain, the family is not consulted nor intervenes in the decision. The remaining participants are divided among those who think (in accordance with current law) that the family conveys what it believes the deceased would have wished and those who think (as actually occurs in practice) that the family can also decide according to its own preferences. When informed that, in Spain, if the deceased’s wishes are not known, the family is allowed to decide, 23% of those surveyed approve of this practice and 56% are ambivalent on this point (Table 3b).

When allocating vital organs, respondents consider utility of transplantation (defined as two times the increment in recipient life expectancy following transplantation) to be less important than other criteria, such as the lesser likelihood of obtaining a suitable organ, urgency, time on waiting list, or the age of the recipient (Table 4).

**Table 4:**
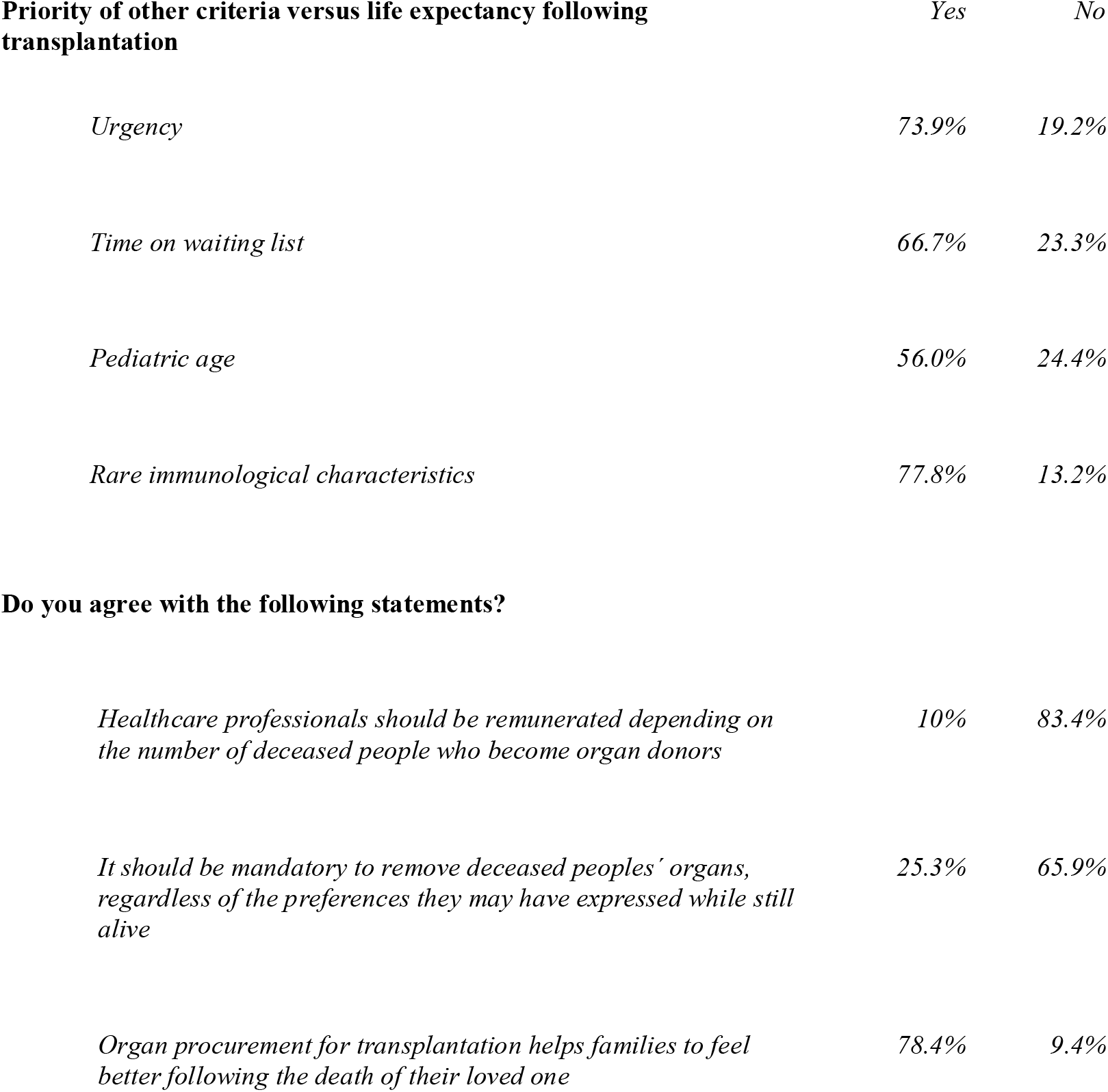
Priority of other allocation criteria over utility of transplantation and Economic incentives, organ conscription and family grieving.

| **Priority of other criteria versus life expectancy following transplantation** | *Yes* | *No* |
| --- | --- | --- |
| *Urgency* | *73.9%* | *19.2%* |
| *Time on waiting list* | *66.7%* | *23.3%* |
| *Pediatric age* | *56.0%* | *24.4%* |
| *Rare immunological characteristics* | *77.8%* | *13.2%* |
| **Do you agree with the following statements?** |  |  |
| *Healthcare professionals should be remunerated depending on the number of deceased people who become organ donors* | *10%* | *83.4%* |
| *It should be mandatory to remove deceased peoples' organs, regardless of the preferences they may have expressed while still alive* | *25.3%* | *65.9%* |
| *Organ procurement for transplantation helps families to feel better following the death of their loved one* | *78.4%* | *9.4%* |

Most participants reject both healthcare personnel involved in the organ recovery process being paid according to the number of deceased people who become organ donors, and the option of implementing an automatic organ recovery model, also known as a “confiscatory” or “conscription” model. Finally, regarding the impact of donation on family grieving, a majority of participants believe that organ recovery for transplantation helps family members to feel better following the death of their loved one (Table 4).

## Discussion

Our study is subject to certain limitations related to the administering method (mixed: online and via telephone) and the sample (a representative panel of the Andalusian population, which might not be extrapolated to the Spanish population as a whole). Given the generally lowering response rates to surveys and increasing costs associated with obtaining samples that meet appropriate quality standards, it is ever more common to resort to online surveys, either alone or combined with other data collection modes. The creation of the online survey was accompanied by a great deal of methodological work that analyzes whether or not the survey’s results are comparable to the findings obtained through other procedures.[14] Based on the conclusions drawn from such literature, one of the most straightforward findings is that, when faced with questions on a sensitive topic, the self-administered modes usually provide more precise answers than those involving an interviewer.[15] Besides, the social desirability bias may be more pronounced in panel-based onIine surveys due to the fact that anonymity is lost from the moment we address participants by name. Given that this study deals with organ donation, social desirability and altruism bias may slant our results toward more favorable attitudes on donation (for example, a stated willingness to donate at a higher than average rate). When analyzing whether or not participants who decide to donate the 5 euros tend to express greater willingness to donate than those who keep them, we have ascertained that this is not the case, which would suggest that social desirability, in this case, does not affect altruism.

Overall, our study shows mostly favorable attitudes towards organ donation in Southern Spain and high levels of trust in the transplant system. This is reflected through a high stated intent to donate one’s own organs after death and to authorize organ removal for a family member. Our study also reveals a certain degree of societal misunderstanding regarding the presumed consent model currently in force in Spain and the clinical criteria for declaring death. Conversely, a majority is aware of the decisive role families play in organ donation and agrees with such a role, which may mitigate other doubts regarding donation procedures. Further research on the social perception of organ donation policies may enable policy makers to better assess opportunities to foster public support of organ donation, and to promote socially acceptable organ procurement policies.

With regard to attitudes toward donation, Spain is slightly above European averages on willingness to donate one’s own organs (61% vs 55%) and authorization for the recovery of a relative’s organs (59% vs 53%).[16] According to national studies, stated willingness to donate in Spain in 2003 and 2011 stood between 63% and 67%.[11,13] These figures were higher in Andalusia, where 86% of the population expressed willingness to donate.[17] Our survey yield intermediate values, with 76% of Andalusians stating their willingness to donate.

In 1993, 1999 and 2006, around half of Spaniards indicated they were willing to authorize organ retrieval for a family member without knowing the latter’s wishes on the subject,[12] while a 2002 study on the Andalusian population raised this proportion to 71% of those surveyed.[18] Our regional study also shows that 62% of Andalusians would authorize organ recovery for a relative whose wishes were unknown, as opposed to 24% who wouldn’t. In practice, the actual rate of family authorization is even higher: 86% in Spain and 89% in Andalusia.[17] Although these figures include both cases (deceased’s willingness to donate and unknown wishes), the high conversion rates –from potential donors to real donors– observed in Spain may be associated with coordination teams’ training and skills in building trust with families along the donation process.[19]

High donation and low family opposition figures may also be explained by Spaniards’ level of trust in their transplantation system. This study reveals respondents high level of trust in the public healthcare system (79%) and transplantation system (93%). European data on the public’s rating of the overall healthcare system indicates that, on a scale of 1 (“very poor”) to 10 (“very good”), the average rating in European countries in 2008 was 4.75, while in Spain it was 6.12 and in Andalusia it was 6.28.[20] In 2016, the rating of the healthcare system in Spain continued to be higher than in European and non-European countries.^30^

Spaniards’ high confidence level in their transplantation system contrasts with their awareness of it. Indeed, over half of those surveyed consider themselves to be somewhat or completely uninformed on the requirements needed to be donors. The widespread lack of awareness regarding Spain’s consent model has been reported in other presumed consent countries.[21] This may impede autonomous decision-making and the fulfillment of peoples’ posthumous wishes concerning organ retrieval, especially among the minority who oppose the removal of their organs after death. In Spain, all adults are considered to be organ donors unless they had expressed their refusal in life. However, 60%) erroneously believe that they would not be considered as donors unless they explicitly say so, and 12% acknowledge not knowing the default policy on this issue. The risk of procuring organs from people opposed to donation only affects a minority –9% according to our survey– which asserts its opposition to donation. Notwithstanding, most of this subgroup (84%) did not express its opposition in any way. In fact, the risk of frustrating individuals’ non-expressed refusal to donate may decrease as a result of involving the family in the decision-making process.

A perhaps relevant difference between opt-in and opt-out jurisdictions is that citizens living in presumed consent systems tend to be less aware of their model of consent for organ donation than citizens in opt-in countries.[21] This study suggests that lack of awareness about the presumed consent system in force may account for the failure of those who reject organ donation to explicitly express their refusal. However, lack of knowledge on the opt-out model may be attributed to the fact that presumed consent is not applied de facto in Spain. Indeed, in practice, transplant coordinators do not proceed with organ retrieval unless the deceased’s family previously authorizes it. Therefore, family members end up having the last word: they may authorize organ recovery in the absence of the deceased’s stated wishes, and they may even oppose recovery when the deceased had expressed their wish to be a donor.[22,23] Reassuringly, our results suggest most people are familiar with this situation: only one out of ten Andalusians erroneously believes that the family plays no role in the decision regarding recovery.

When directly surveyed on their opinion regarding the current model of presumed consent, participants in our survey display less opposed attitudes than in other studies. In preliminary surveys conducted in Southern Spain, between 65% and 75% of participants opposed presumed consent.[13,24,25] In our study, 30% of those surveyed expressed their opposition to this model (once briefed on the opt-out model currently in force), while 44% asserted that they neither agree nor disagree with the model. Furthermore, when offered various options, the proportion of those who preferred presumed consent (40%) was comparable to the proportion of the surveyed who preferred the explicit consent model (44%). Most of the Andalusian population are not only familiar with the key role afforded to families in the decision-making process for organ recovery, but also approve of them having such a key function. The fact that only a minority (19%) disapprove of family members making that decision suggests that our participants mainly trust the criteria of their loved ones to uphold and defend their posthumous interests. We venture that the high degree of confidence that Spanish society places upon family bonds may end up reducing other concerns regarding organ recovery practices –for example, those expressed with regard to death diagnosis– thereby neutralizing any related objections.

Given widespread practices of donation after brain death and DCD, and the fact that the public displays a generic predisposition favorable to donation, it is still surprising that three out of four participants refuse donation, or doubt whether or not to donate, when presented with a schematic description of the prototypical clinical situations in which cadaveric donation actually takes place: brain death, controlled DCD and uncontrolled DCD. This finding, which apparently contradicts the overall willingness to donate, may suggest that support for donation amongst the surveyed population is somewhat superficial: although favorable to donation, it runs into uncertainty when it becomes aware of the details of the specific clinical circumstances in which donation actually takes place. Alternately, this finding may signal that participants have erroneously assumed that the scenarios described clinical situations where candidates for organ recovery were still alive. In spite of the fact that the description of each case reflected the fulfillment of the legally established criteria for being declared dead, the words “death” and “dead” were deliberately omitted. In support of this second hypothesis, we should highlight that the majority did not acknowledge these situations as clinical scenarios in which organ recovery is legal.

The ethical problems associated with organ allocation for transplantation are usually explained as a compromise between transplant utility -understood as the optimization of this resource, and measured in terms of recipient’s life expectancy-[26] and other competing non-utilitarian criteria, such as urgency, time on waiting list, pediatric age, and rare immunological characteristics. A recent systematic review on general public preferences[27] shows that the preferred organ allocation criteria is recipient’s life expectancy corrected for urgency, which the authors call “the ethical rational utilitarian model” and which accounts for urgency. Another systematic review also encounters a preference for utilitarian criteria among healthcare professionals, which is not the case among patients, who mostly lean toward urgency.[28] Our study shows that a majority of the public prefers allocating organs based on non-utilitarian criteria, which is consistent with other surveys on allocation conducted over the last two decades within diverse segments of the population and in different countries.[29,30] Further research is needed to better understand lay people’s preferred moral pathways in organ allocation.

## Conclusions

This study shows attitudes within the Southern population in Spain mostly favorable to organ donation and the transplantation system. Most of those surveyed trust their transplant system, and this is reflected through a high stated intent to donate their organs after death and to authorize organ removal for a family member, even when the latter’s preferences on the subject are unknown. Our study also reveals a certain degree of societal misunderstanding regarding the presumed consent model currently in force in Spain, and regarding the legality of procuring organs from candidates for donation who fulfill the clinical criteria for either brain death or circulatory death. Conversely, a majority is aware that, in Spain, families of the deceased actually play a decisive role, and most of those surveyed do not express opposition to families’ being given such a key function. Awareness that families perform this key role in final decision-making may explain why so few Spaniards, including those who refuse to donate, express their preferences in written form. It may also reduce or mitigate other doubts or ethical objections that the population expresses regarding donation procedures, such as those related to presumed consent and ambivalence regarding the criteria for determining donors’ death. Further research on the social perception of potentially controversial organ donation policies may enable policy makers to better assess opportunities to foster public support of organ donation, and to anticipate possible threats to the trust citizens place in donation and transplantation policies.

## Data Availability

Information about the methodology can be access through the link below and the reference is: PACIS EP-1807 IESA-CSIC.

http://www.iesa.csic.es/pacis/

## Abbreviations

DCD: donation after circulatory death
PACIS: Citizens’ Panel for Social Research in Andalusia

## Authorship

All co-authors have made substantial contributions to this manuscript and their participation warrants authorship. Specific contributions are detailed below.

- **Gonzalo Díaz-Cobacho:** Participated in the writing of the original draft, and coordinated the investigation
- **Maite Cruz-Piqueras:** Participated in data analysis and in the writing of the manuscript
- **Janet Delgado:** Participated in data analysis and in the writing of the manuscript
- **Joaquín Hortal-Carmona:** Participated in data analysis and in the writing of the manuscript
- **M. Victoria Martínez-López:** Participated in data analysis and in the writing of the manuscript
- **Alberto Molina-Pérez:** Participated in research design, performance of the research, in the writing the original draft, and project administration,
- **Alvaro Padilla-Pozo:** Coordinated the data analysis, and participated in the writing of the manuscript.
- **Julia Ranchal-Romero:** Participated in the research design, coordinated sampling and data gathering, methodology, formal analysis, and the writing of the manuscript.
- **David Rodríguez-Arias:** Participated in research design, performance of the research, writing of the original draft, and project administration.

## Acknowledgments

We would like to thank Jose Luis Rodriguez-Arias for assistance with the statistical analysis, Beatriz Domínguez-Gil and Alicia Pérez Blanco for helpful suggestions that have improved the original manuscript, and Scott A. Singer for translation assistance.

## Tables

In the tables that we will show below, the percentages do not add up to 100% because we have not included the non-responders.

## Notes

***Conflict of interest disclosure:*** The authors declare no conflicts of interest.

### Competing Interest Statement

The authors have declared no competing interest.

### Funding Statement

This research was carried out through Project INEDyTO [Research on the Ethics of Organ Donation and Transplantation], funded by the Spanish Government (MINECO FFI2017-88913-P) in collaboration with the Institute for Advanced Social Studies (IESA-CSIC), which sponsored the data collection and participated in the analysis and interpretation of data.

### Author Declarations

The study protocol was reviewed by the Coordinating Committee on Biomedical Research Ethics of Andalusia (PEIBA 2521-N-20), which waived full review for this type of study. Written informed consent was obtained from all individual participants prior to participation.

## References

[1] Iacobucci G. Proposals for opt-out organ donation launched for England. BMJ 2017;359:57–64. https://doi.org/10.1136/bmj.j5764.

[2] Matesanz R, Domínguez-Gil B, Coll E, de la Rosa G, Marazuela R. Spanish experience as a leading country: what kind of measures were taken?: Facing organ shortage in Spain. Transplant International 2011;24:333–43. https://doi.org/10.1111/j.1432-2277.2010.01204.x.

[3] Wakefield MA, Loken B, Hornik RC. Use of mass media campaigns to change health behaviour. The Lancet 2010;376:1261–71.

[4] Delmonico FL, Arnold R, Scheper-Hughes N, Siminoff LA, Kahn J, Youngner SJ. Ethical incentives - Not Payment - For Organ Donation. The New England Journal of Medicine 2002;346. https://doi.org/10.1056/NEJMsb013216.

[5] Manyalich M, Mestres CA, Ballesté C, Páez G, Valero R, Gómez MP. Organ procurement: Spanish transplant procurement management. Asian Cardiovasc Thorac Ann 2011;19:268–78. https://doi.org/10.1177/0218492311411590.

[6] EDQM. Safety, quality and ethical matters related to the use of organs, tissues and cells of human origin. Strasbourg: European Directorate for the Quality of Medicines and HealthCare; 2017.

[7] Organización Nacional de Trasplantes. Actividad de donación y trasplante España 2019. Madrid: Organización Nacional de Trasplantes; 2020.

[8] Caballero F, Matesanz R. Manual de Donación y Trasplante de Órganos Humanos. Ministerio de Sanidad, Servicios Sociales e Igualdad; 2015.

[9] Matesanz R. El modelo español de coordinación y trasplantes. Madrid: Aula Médica; 2008.

[10] Conesa C, Ríos A, Ramírez P, Canteras M, Rodríguez MM, Parrilla P. [Multivariate study of the psychosocial factors affecting public attitude towards organ donation]. Nefrologia 2005;25:684–97.

[11] Conesa C, Ríos A, Ramírez P, Rodríguez MM, Rivas P, Canteras M, et al. Psychosocial profile in favor of organ donation. Transplantation Proceedings 2003;35:1276–81. https://doi.org/10.1016/S0041-1345(03)00468-8.

[12] Domínguez-Gil B, Martín MJ, Valentín MO, Scandroglio B, Coll E, López JS, et al. Decrease in refusals to donate in Spain despite no substantial change in the population’s attitude towards donation. Organs, Tissues & Cells 2010;13:17–24.

[13] Scandroglio B, Domínguez-Gil B, Lopez J soler, Valentín MO, Martín MJ, Coll E, et al. Analysis of the attitudes and motivations of the Spanish population towards organ donation after death. Transplant International□: Official Journal of the European Society for Organ Transplantation 2011;24:158–66. https://doi.org/10.1111/j.1432-2277.2010.01174.x.

[14] Ansolabehere S, Schaffner B. Does Survey Mode Still Matter? Findings From a 2010 Multi-Mode Comparison. Political Analysis 2014;22:285–303.

[15] Tourangeau R, Conrad FG, Couper MP. The science of web surveys. Oxford University Press; 2013.

[16] European Commission. Special Eurobarometer 333a: Organ donation and transplantation. Brussels: 2010.

[17] Junta de Andalucía. Nueve de cada 10 andaluces dicen “sí” a la donación de órganos durante 2019. Noticias de La Junta 2020. http://www.juntadeandalucia.es/presidencia/portavoz/salud/149144/ConsejeriaSaludFamilias/donacion/organos/trasplantes (accessed January 15, 2020).

[18] Rando Calvo B, Blanca MJ, de Frutos Sanz MA. La toma de decisión sobre donación de órganos en la población andaluza. Psicothema 2002;14:300–9.

[19] Gómez P, Santiago C, Getino A, Moñino A, Richart M, Cabrero J. La entrevista familiar: Enseñanza de las técnicas de comunicación. Nefrología 2001;21:57–64.

[20] European Social Survey. ESS Round 4: European Social Survey Round 4 Data. Norway: Norwegian Centre for Research Data; 2008.

[21] Molina-Pérez A, Rodríguez-Arias D, Delgado-Rodríguez J, Morgan M, Frunza M, Randhawa G, et al. Public knowledge and attitudes towards consent policies for organ donation in Europe. A systematic review. Transplant Rev 2019;33:1–8. https://doi.org/10.1016/j.trre.2018.09.001.

[22] Delgado J, Molina-Pérez A, Shaw D, Rodríguez-Arias D. The Role of the Family in Deceased Organ Procurement. A Guide for Clinicians and Policy Makers. Transplantation 2019;103:e112–8. https://doi.org/10.1097/TP.0000000000002622.

[23] Rosenblum AM, Horvat LD, Siminoff LA, Prakash V, Beitel J, Garg AX. The authority of next-of-kin in explicit and presumed consent systems for deceased organ donation: an analysis of 54 nations. Nephrol Dial Transplant 2012;27:2533–46. https://doi.org/10.1093/ndt/gfr619.

[24] Martínez-Alarcón L, Ríos A, Sánchez J, Ramis G, López-Navas A, Ramírez P, et al. Evaluation of the law of presumed consent after brain death by Spanish journalism students. Transplant Proc 2010;42:3109–12. https://doi.org/10.1016/j.transproceed.2010.05.073.

[25] Conesa C, Ríos Zambudio A, Ramírez Romero P, Rodríguez Martínez MM, Canteras Jordana M, Parrilla Paricio P. Actitud de la población hacia una legislación de consentimiento presunto a la donación de órganos de cadáver. Medicina clinica 2004;122:67–9.

[26] Freeman RB, Bernat JL. Ethical Issues in Organ Transplantation. Progress in Cardiovascular Diseases 2012;55:282–9. https://doi.org/10.1016/j.pcad.2012.08.005.

[27] Oedingen C, Bartling T, Mühlbacher AC, Schrem H, Krauth C. Systematic Review of Public Preferences for the Allocation of Donor Organs for Transplantation: Principles of Distributive Justice. Patient 2019;12:475–89. https://doi.org/10.1007/s40271-019-00363-0.

[28] Bartling T, Oedingen C, Kohlmann T, Schrem H, Krauth C. Comparing preferences of physicians and patients regarding the allocation of donor organs: A systematic review. Transplantation Reviews 2020;34:100515. https://doi.org/10.1016/j.trre.2019.100515.

[29] Duarte PS, Pericoco S, Miyazaki MCOS, Ramalho HJ, Abbud-Filho M. Brazilian’s Attitudes Toward Organ Donation and Transplantation. Transplantation Proceedings 2002;34:458–9. https://doi.org/10.1016/s0041-1345(02)02595-2.

[30] Geddes CC, Rodger SC, Smith C, Ganai A. Allocation of Deceased Donor Kidneys for Transplantation: Opinions of Patients With CKD. American Journal of Kidney Diseases 2005;46:949–56. https://doi.org/10.1053/j.ajkd.2005.07.031.

